# Why Clinical Trials Stop: The Role of Genetics

**DOI:** 10.1101/2023.02.07.23285407

**Authors:** Olesya Razuvayevskaya, Irene Lopez, Ian Dunham, David Ochoa

**Affiliations:** Open Targets, Wellcome Genome Campus, Hinxton, Cambridgeshire CB10 1SD, UK; European Molecular Biology Laboratory, European Bioinformatics Institute (EMBL-EBI), Wellcome Genome Campus, Hinxton, Cambridgeshire CB10 1SD, UK; Wellcome Sanger Institute, Wellcome Genome Campus, Hinxton, Cambridgeshire CB10 1SA, UK

## Abstract

Many drug discovery projects are started, but few progress fully through clinical trials to approval. Previous work has shown that human genetics support for the therapeutic hypothesis increases the chance of trial progression. Here, we applied natural language processing to classify the freetext reasons for 28,842 clinical trials that stopped before their endpoints were met. We then evaluated these classes in the light of the underlying evidence for the therapeutic hypothesis and target properties. We show that trials are more likely to stop due to lack of efficacy in the absence of strong genetic evidence from human populations or genetically-modified animal models. Furthermore, trials are more likely to stop for safety reasons if the drug target gene is highly constrained in human populations and if the gene is not selectively expressed. These results support the growing use of human genetics to evaluate targets for drug discovery programmes.

## Introduction

In drug discovery, many projects are started but few yield approved drugs. While there are myriad possible reasons for individual failures in clinical trials, major, and expensive, sources of attrition (79%) are lack of efficacy or unforeseen safety issues, both suggesting that the therapeutic hypothesis was insufficiently supported (Dowden and Munro, 2019). New approaches adopted across the industry aim to improve success rates by systematic assessment of the available evidence throughout the research and clinical pipelines (Morgan et al., 2018; Wu et al., 2021). Recently, support from genetic studies has been associated with successful clinical trial progression (Nelson et al., 2015; King, Davis and Degner, 2019; Ochoa et al., 2022). However further understanding of the reasons for success or failure in clinical trials could assist in reducing future attrition.

Systematically assessing the reasons for success or failure in clinical trials is hampered by several factors. Several surveys have demonstrated a bias towards reporting positive results, with 78.3% of trials in the literature reporting successful outcomes (Ioannidis, 2005; Young, Ioannidis and Al-Ubaydli, 2008; Bourgeois, Murthy and Mandl, 2010). Successful clinical trials are published significantly faster than trials reporting negative results (Qunaj *et al*., 2018; Jones *et al*., 2021). Clearly, there is a disconnect between reporting of positive trial results and overall success rates (DiMasi *et al*., 2010). Having access to negative results is important to reveal efficacy tendencies and safety liabilities (Petsko, 2010), for retrospective review and for benchmarking of predictive methods including machine learning (ML). One area that has not been examined extensively is the reasons underlying the premature stopping of clinical trials. Many clinical trials stop before their scheduled endpoint is met and before formal success or failure outcomes are declared.

ClinicalTrials.gov is a free-to-access database registering global clinical trials (Ross *et al*., 2009; Califf *et al*., 2012) and provides a partially structured trial status. For trials halted before their scheduled endpoint, ClinicalTrials.gov provides a freeform stopping reason - termination, suspension or withdrawal (Al-Durra *et al*., 2018). Pak *et al*. previously classified stopping reasons for 3,125 stopped trials and found that only 10.8% of trials stopped because of a clear negative outcome, while the majority (54.5%) fell into a set of reasons characterised as neutral (Pak, Rodriguez and Roth, 2015). Here, we extended that work by training a natural language processing (NLP) model to classify stopping reasons and used this model to classify 28,842 stopped trials. We integrated our classification with evidence associating the drug target and disease from the Open Targets Platform (Ochoa *et al*., 2023), revealing that trials stopped for lack of efficacy or safety reasons were less supported by genetic evidence. Furthermore, trials involving drugs whose target gene is constrained in human populations were more likely to stop for safety reasons (Duffy *et al*., 2020), while drugs with targets with tissue-selective expression were less likely to pose safety risks. Taken together these observations confirm and extend previous studies recognising the value of genetic information and selective expression in target selection.

## Results

### Interpretable classification of the reasons why clinical trials stop

To catalogue the reasons behind the withdrawal, termination or suspension of clinical studies, we classified every free-text reason submitted to ClinicalTrials.gov using an NLP classifier. In order to build a training set for our model, we revisited the manual classification of 3,124 stopped trials based on the available submissions to ClinicalTrials.gov in May 2010 (Pak, Rodriguez and Roth, 2015). The authors classified every study with a maximum of 3 classes following an ontological structure (Supplementary Table 1). Each of the classes was also assigned a higher-level category representing the outcome implications for the clinical project. For example, 33.7% of the studies were classified as stopped due to *“*insufficient enrollment*”*, a *“*neutral*”* outcome due to its expected independence from the therapeutic hypothesis. When inspecting submitted reasons belonging to the same curated category, we observed a strong linguistic similarity, as revealed by clustering the cosine similarity of the sentence embeddings (Supplementary Figure 1). Studies stopped because of reasons linked to lack of efficacy and studies stopped due to futility have a linguistic similarity of 0.98, with both classes manually classified as *“*negative*”* outcomes. Based on this clustering, we redefined the classification by merging semantically similar classes represented by low numbers of annotated sentences. Moreover, we added 447 studies stopped due to the COVID-19 pandemic (Supplementary Table 2), resulting in a total of 3,571 studies manually classified into at least one of 17 stop reasons and explained by 6 different higher-level outcome categories.

By leveraging the consistent language used by the submitters, we fine-tuned the BERT model (Devlin *et al*., 2018) for the task of clinical trial classification into stop reasons (Supplementary Methods). Overall, the model showed strong predictive power in the cross-validated set (*F*_*micro*_=0.91), performing strongly for the most frequent classes, such as *“*insufficient enrollment*”* (F =0.98) or *“*COVID-19*”* (F=1.00), but demonstrating decreased performance on linguistically complex reasons, such as trials stopped because of another study (F=0.71) (Supplementary Table 3).

To further evaluate the model, we manually curated an additional set of 1,675 stop reasons from randomly selected studies not included in the training set. Overall, the performance against the unseen data was lower but comparable to that of the cross-validated model (*F* _*micro*_ ranging from 0.70 to 0.83 depending on the choice of the annotator) (Supplementary Table 4), demonstrating real-world performance and reduced risk of overfitting. Interestingly, the curators demonstrated a relatively low agreement for many classes where the ML model also showed relatively weak performance, such as studies stopped due to insufficient data or met endpoint (Supplementary Methods, Supplementary Figure 12).

### Stop reasons reflect operational, clinical and biological constraints

Classification of the 28,842 stopped trials submitted to ClinicalTrials.gov before 27/11/2021 was performed using our NLP model fine-tuned on all the manually curated sentences (Supplementary Table 5). 99% of the trials were classified with at least 1 of the 15 potential reasons and mapped to one of 6 different higher-level outcomes (Figure 1). *“*Insufficient enrollment*”* remained the most common reason to stop a trial (33.52%), with other reasons prior to the accrual of any study results also occurring in a large number of studies. A total of 977 trials (3.38%) were classified as stopped due to *“*Safety or side effects*”* and 2,197 studies (7.6%) were stopped due to negative reasons, such as those questioning the efficacy or value (futility). The relative frequency of each stop reason varies across clinical phases, reflecting the purposes of each study (Supplementary Figure 2). For example, studies with negative outcomes were more frequently reported during Phase II and Phase III, whereas safety or side effect outcomes were proportionally more frequent in Phase I. Trials stopped due to the relocation of the study or key staff were instead more frequent in Early Phase I. Of all the studies with a submitted stop reason, 48% were indicated for oncology, an expected overrepresentation due to the recent inclusion of the stop reasons in ClinicalTrials.gov. Moreover, oncology studies stopped more frequently due to safety or side effects and were rarely stopped due to the COVID-19 pandemic (Supplementary Figure 3). Alternatively, COVID-19 was the reported reason to stop respiratory studies at a higher rate than any other therapeutic area, possibly indicating increased operational difficulties.

**Figure 1:**
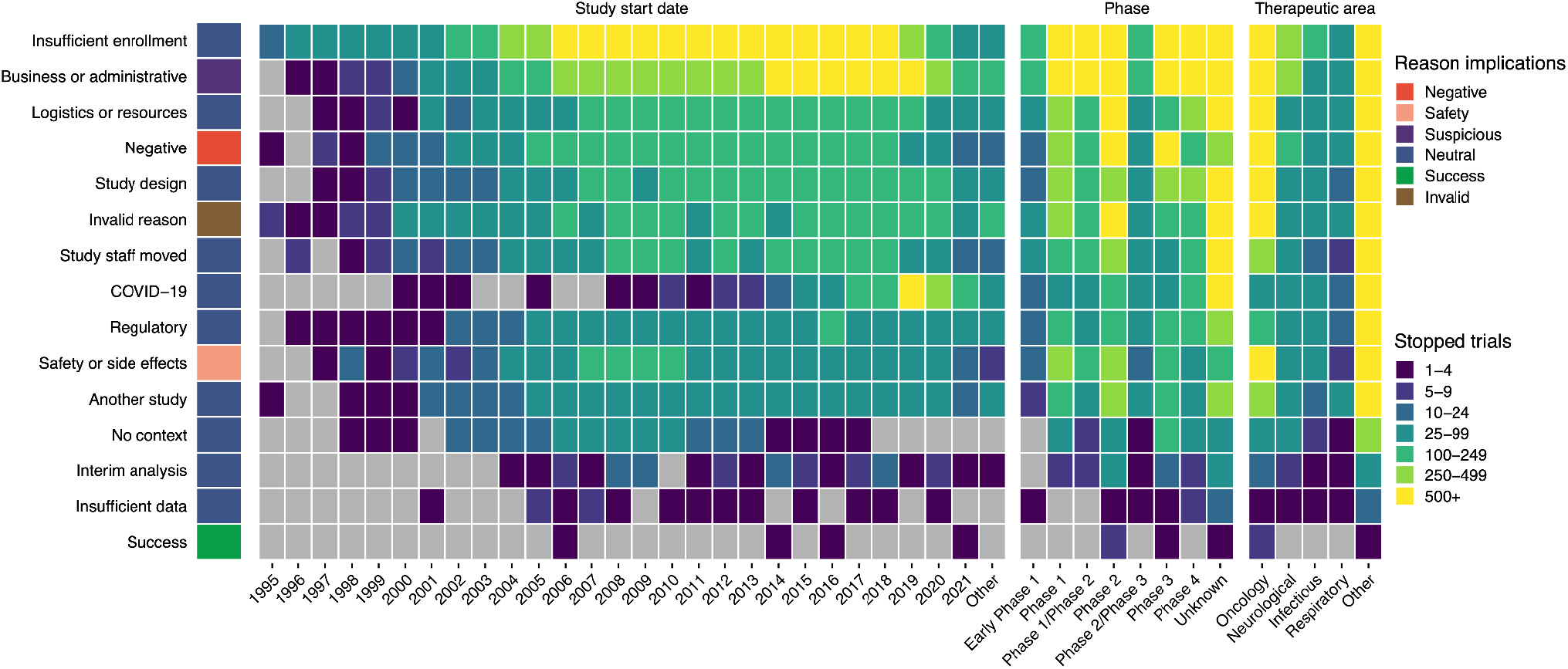
Classification of stopped reasons for 28,842 clinical trials in ClinicalTrials.gov. Predicted trial stop reasons are shown in rows with counts of trials per start year, clinical phase or therapeutic area shown by the colour in each cell. The outcome groupings of the stopped reasons are shown by means of the colour next to the stopped reason label. Note that trials start potentially many years before they are stopped.

### Availability of genetic support for stopped trials influences the outcome

To better understand the underlying reasons that might have caused the study to fail, we assessed the availability of different types of potentially causal genetic evidence for the intended pharmacological targets in the same indication. By using genetic evidence collated by the Open Targets Platform, we reproduced previous reports indicating that genetically supported studies are more likely to progress through the clinical pipeline (Figure 2) (Nelson *et al*., 2015; King, Davis and Degner, 2019). Interestingly, we also observed that stopped trials - among all the trials at any phase - are depleted in genetic support (OR =0.75, p = 6e-60). A similar lack of genetic evidence was observed for the 3 types of stopped studies: withdrawn, terminated and suspended (Supplementary Table 6).

**Figure 2.**
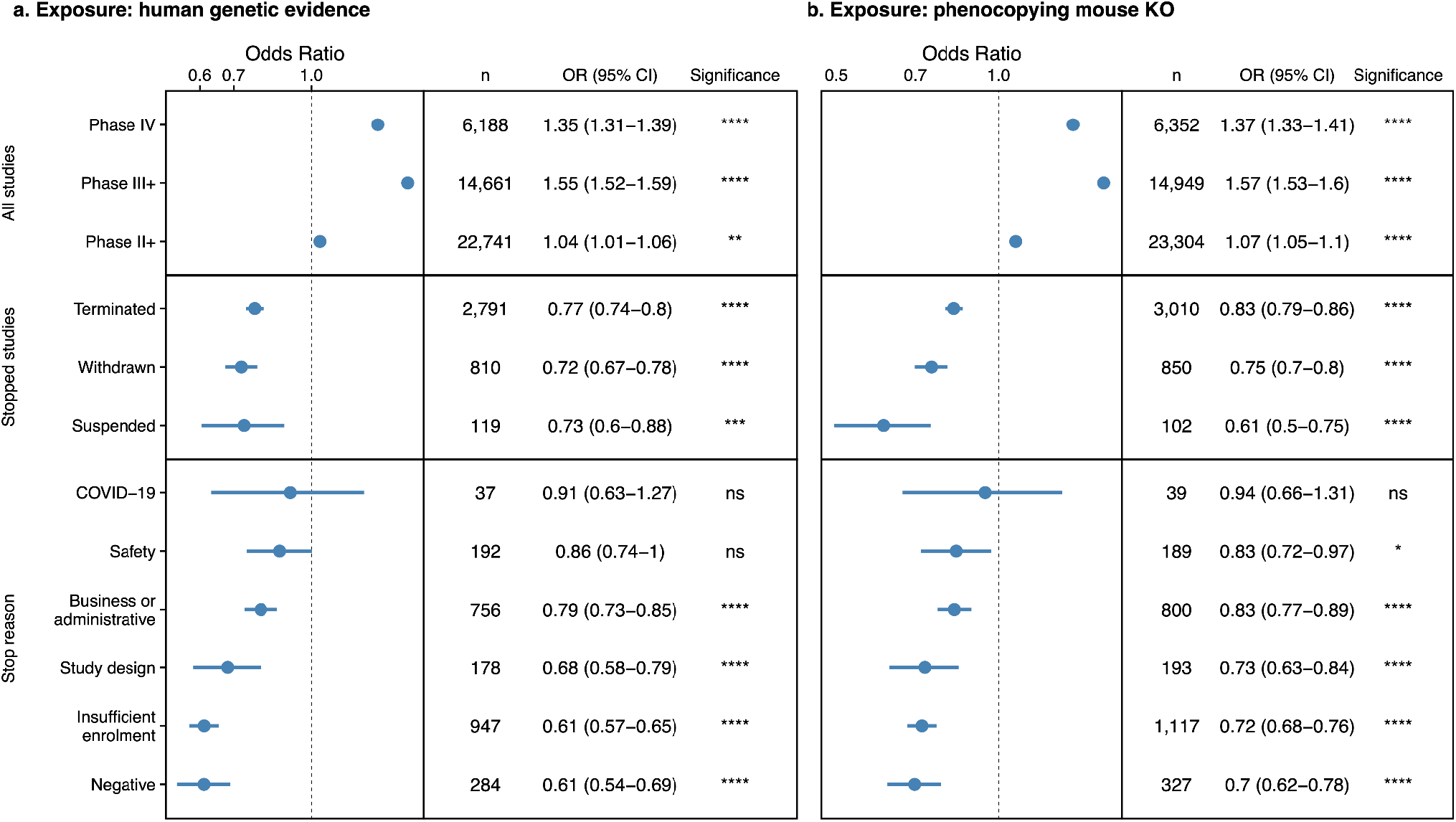
Association between the availability of genetic evidence and clinical trial outcomes. Genetic evidence support for clinical trials either from human genetics studies (panel a) or IMPC mouse knockouts that phenocopy the human disease (panel b). The panels show the odds ratio of support for the target-disease hypothesis from genetics evidence for all clinical trials split by phase (top row), stopped clinical trials (centre row) and stopped clinical trials split by higher level stopping reason (bottom row). Significant odds ratios > 1 are enriched and <1 are depleted for genetic evidence in each subclass of trial. Significance levels: **** *p* < 1e-4; \*\*\**p* < 1e-3; ** *p* < 1e-2; * *p* < 0.05; *“*ns*”* not significant.

When stratifying the stopped studies by reason, trials halted due to negative outcomes - such as lack of efficacy or futility - displayed a significant decrease of genetic support for the intended pharmacological target in the same indication (OR = 0.61, p = 6e-18) (Figure 2). The depletion of evidence on negative outcomes remains consistent when looking at different sources of genetic evidence in common or rare diseases including genome-wide association studies (GWAS) processed by the Open Targets Genetics Portal (Ghoussaini *et al*., 2021), gene burden tests based on sequencing of large population cohorts (Backman *et al*., 2021; Wang *et al*., 2021; Karczewski *et al*., 2022), ClinVar (Landrum *et al*., 2014), ClinGen Gene Validity (McGlaughon *et al*., 2018), Genomics England PanelApp (Baker, 2018), gene2phenotype (Thormann *et al*., 2019), Orphanet (Rodwell and Aymé, 2015), and Uniprot (UniProt Consortium, 2019) (Supplementary Figure 4).

Other predicted reasons for stopping the trials such as insufficient enrolment, problems with the study design or business or administrative reasons also present a strong to moderate depletion of genetic evidence denoting potential reduced support for the therapeutic hypothesis (Figure 2). We found studies stopped due to coincidental factors such as the COVID-19 pandemic or safety and side effects have no association with the availability of genetic support for the intended target in the primary indication.

The observed associations between clinical trial outcomes and the availability of genetic support remain consistent when considering genetic information in mouse models (Figure 2). Stopped trials due to negative factors present the weakest support among all predicted reasons (OR = 0.7, p = 4e-11) when genetic evidence is defined as the presence of a murine model in which the drug target homologous gene knockout causes a phenotype that mimics the indication as reported by the IMPC (Muñoz-Fuentes *et al*., 2018).

### Genetically constrained targets anticipate safety-associated stopped trials

Analysis of the classified stop reasons indicates that oncology trials are 2.4 times more likely to stop because of safety or side effects (OR = 2.14, p = 8.1e-79, Figure 3). Moreover, for all trials predicted to stop due to safety concerns we found a significant enrichment in targets associated with driver events reported by COSMIC (Bamford *et al*., 2004), ClinVar (Landrum *et al*., 2014) or IntOgen (Gundem *et al*., 2010) (Supplementary Figure 5). Examining the target properties, we found that studies targeting genes highly constrained in natural populations (GnomAD pLOEUF 16th percentile) are 1.5 times more likely to stop due to safety concerns (Karczewski and Francioli, 2017). Furthermore, the risk of stopping because of safety declines as the genetic constraint of the target decreases. Similarly, we identified a 1.4-fold increased risk of stopping due to safety concerns when the targeted gene is classified as loss-of-function intolerant (pLi > 0.9). We also identified functional genomic features that inform on increased safety risk. A similar 1.4-fold increased risk is observed for genes expressed with low tissue specificity according to the human protein atlas (Uhlén *et al*., 2015). Instead, studies targeting tissue-enriched genes show a lower-than-expected (OR = 0.83, p = 6.8e-4) likelihood of stopping due to safety. Finally, targets physically interacting with 10 or more different partners according to the IntAct database (mi score > 0.42) present an increased risk of stopping due to safety.

**Figure 3.**
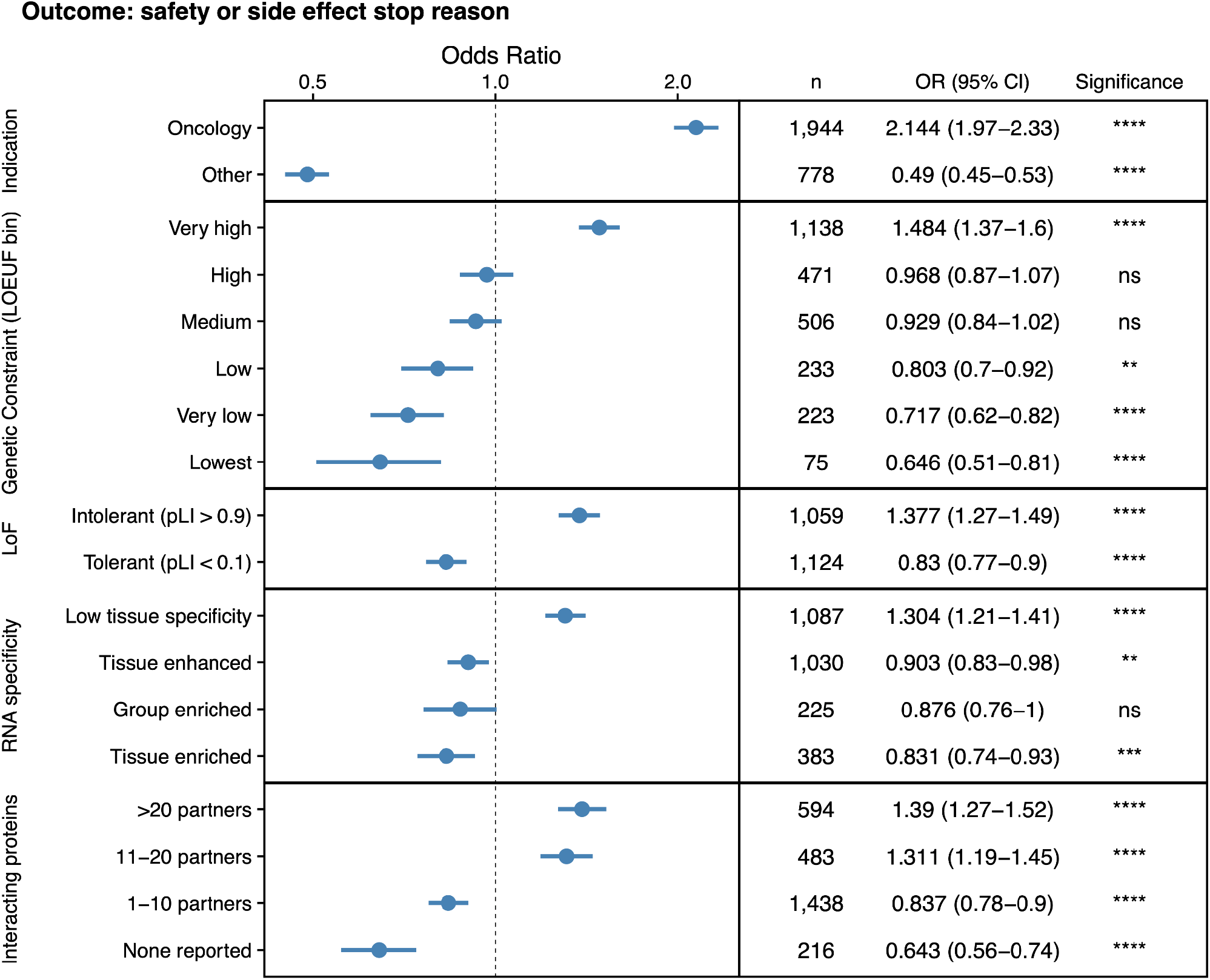
Risk of stopping a clinical trial due to safety or side effects examined by therapeutic area and target properties. Odds ratio (OR) > 1 represents increased risk of study stopping and OR < 1 protection against stopping. Clinical trials are split by therapeutic area (oncology vs others) in the top section, by types of tissue specificity (second section), relative genetic constraint of the target (LOEUF, middle section), Intolerance to loss of function (LOF) mutations (penultimate section) and by network connectivity (bottom section).

## Discussion

Genetic evidence is increasingly leveraged by the pharmaceutical industry to add support to the therapeutic hypothesis (Barrett, Dunham and Birney, 2015; Morgan *et al*., 2018; Wu *et al*., 2021; Fernando *et al*., 2022). Adding to previous observations on the role of genetic factors in the overall trial success (Nelson *et al*., 2015), (Nelson *et al*., 2015; King, Davis and Degner, 2019), we exploited under-utilised data from clinical trial records to better understand the opposite outcome: why clinical trials stop. While the availability of genetic evidence might inform future success, failure remains the most common outcome of clinical studies and - to our knowledge - no systematic evidence exists on the relevance of genetics to de-risk negative results.

Recent reports indicate that 79% of clinical studies fail because of the lack of efficacy or safety (Dowden and Munro, 2019). Our analysis indicates that within the 7.4% of studies that stop early - due to withdrawal, termination or suspension - the proportion of trials that failed because of efficacy or safety is only 10.4%. Stopped studies are more likely to fail due to early coincidental factors not necessarily linked to biological plausibility e.g. the PI relocates or insufficient enrollment in the trial. Notwithstanding the reduced relative risk of efficacy and safety as the main causes for stopping the trial, these studies provide a significant body of unsuccessful results likely explained by a weak therapeutic hypothesis. Continued expansion of the recording of negative results from clinical trials, including stoppages, will be valuable. To assist in this effort, we will continue to update the classification of stopped studies through the Open Targets Platform (https://platform.opentargets.org) (Ochoa *et al*., 2023). Further investigation of the study outcomes for completed studies could expand our understanding of the reasons behind unsuccessful trials, particularly after the accrual of the study results.

Our analysis exploits the classified stop reasons to understand the relative importance of the causes leading to failed studies. By using a case-control approach, we conclude that genetic support is not only predictive of clinical trial progression but also protective of early trial stoppage. We illustrate different ways in which genetic causality and genetic constraint can de-risk the target selection process. However, many stopped trials, even for efficacy and safety reasons, might be explained by factors beyond the intended pharmacological target. Off-target effects, pharmacokinetics, drug delivery or toxicology are other risks not considered in this study that might also explain a set of negative outcomes. Another limitation of our study is that the reasons submitted to ClinicalTrials.gov might only represent a fraction of all the reasons contributing to the decision to halt the study. For example, we found that studies classified as stopped due to patient recruitment manifest weaker genetic support, an observation that we did not anticipate due to the lack of an obvious link between enrollment and biological plausibility. Hence, we hypothesise a fraction of the stopped trials might present an overall lack of confidence in the therapeutic hypothesis, independently of the reported reason.

This study showcases how reflecting on past failures can inform on the relative importance of the risks associated with early target identification and prioritisation. Although clinical trial success is a discrete outcome, failure needs to be understood as a breakdown of many possible causes. A proper set of positive and negative outcomes such as the ones introduced in this work represent the groundwork necessary to implement quantitative or semi-automatic models to objectively de-risk any future studies.

## Data Availability

All data and methods in the present work are available online as described in the respective sections of the manuscript.

## Acknowledgements

We would like to thank Teodore R. Pak, Maria D. Rodriguez, and Frederick P. Roth for providing the dataset of curated stop reasons that was used for training our model. We are grateful to our curators, Andrew Hercules, Irene Lopez, Asier Gonzalez, Konstantinos Tsirigos, and Helena Cornu for volunteering to participate in the annotation study. Finally, we would like to thank Sandra Machlitt-Northen for providing detailed feedback on the re-defined categories for stopped trials.

## Abbreviations

BERT: Bidirectional Encoder Representations from Transformers
ClinGen: The Clinical Genome
ClinVar: Clinically relevant Variants
COSMIC: Catalogue of Somatic Mutations in Cancer
gnomAD: The Genome Aggregation Database
WAS: Genome-Wide Association Studies
IntOgen: Integrative Onco Genomics
LOEUF: Loss-Of-Function Observed/Expected Upper Bound Fraction
LOF: Loss-Of-Function
NLP: Natural Language Processing OR - Odds Ratio
PI: Principal Investigator
UniProt: Universal Protein resource

## Methods

In order to redefine the stop reason ontology provided by Pak *et al*., we trained a Long-Short Term Memory Network (LSTM) to create a representation for each reason and calculated pair-wise cosine similarities of the resulting embeddings after averaging all examples in the same class (Supplementary Methods). Similar classes with low number of sentences were grouped together when accounting for similar reasons (Supplementary Table 1). To train our multiclass multilabel stop reason classifier, we fine-tuned a BERT uncased pre-trained model with a one-layer feed-forward classifier for the task of predicting the termination reasons. Training set (https://huggingface.co/datasets/opentargets/clinical_trial_reason_to_stop) as well as the resulting model (https://huggingface.co/opentargets/stop_reasons_classificator_multi_label) are available in hugging face for interactive inspection.

To collate the genetic support for each clinical trial the bioactive molecules in each study were mapped to their intended pharmacological targets using the mechanism of action curated by the ChEMBL database (Gaulton *et al*., 2017). Genetic support for human and mouse models for the exact indication was extracted from 13 different sources covering rare, common and somatic indications integrated by the Open Targets Platform (Ochoa *et al*., 2023). Significance of each case-control study was computed using a Fisher Exact test on all clinical trials independently of their status or whether they are stopped. Genetic information was propagated to ancestors in the Experimental Factor Ontology to account for differences on curation. Code to reproduce the analysis is available in https://github.com/opentargets/stopReasons.

## Supplementary Tables

**Supplementary Table 1**. *Pak et al*. manual curation of stopped studies.

**Supplementary Table 2**. Curated reasons assigned to be due to COVID-19 pandemic

**Supplementary Table 3**. Model performance on Pak et al. + COVID-19 and tested on Pak et al. + COVID-19

**Supplementary Table 4**. Model performance trained on Pak et al. + Covid-19 data and tested on unseen data by annotator

**Supplementary Table 5**. NLP model classification of stopped studies

**Supplementary Table 6**. Case-control test results for all combinations of exposures and outcomes.

## Supplementary Figures

**Supplementary Figure 1:**
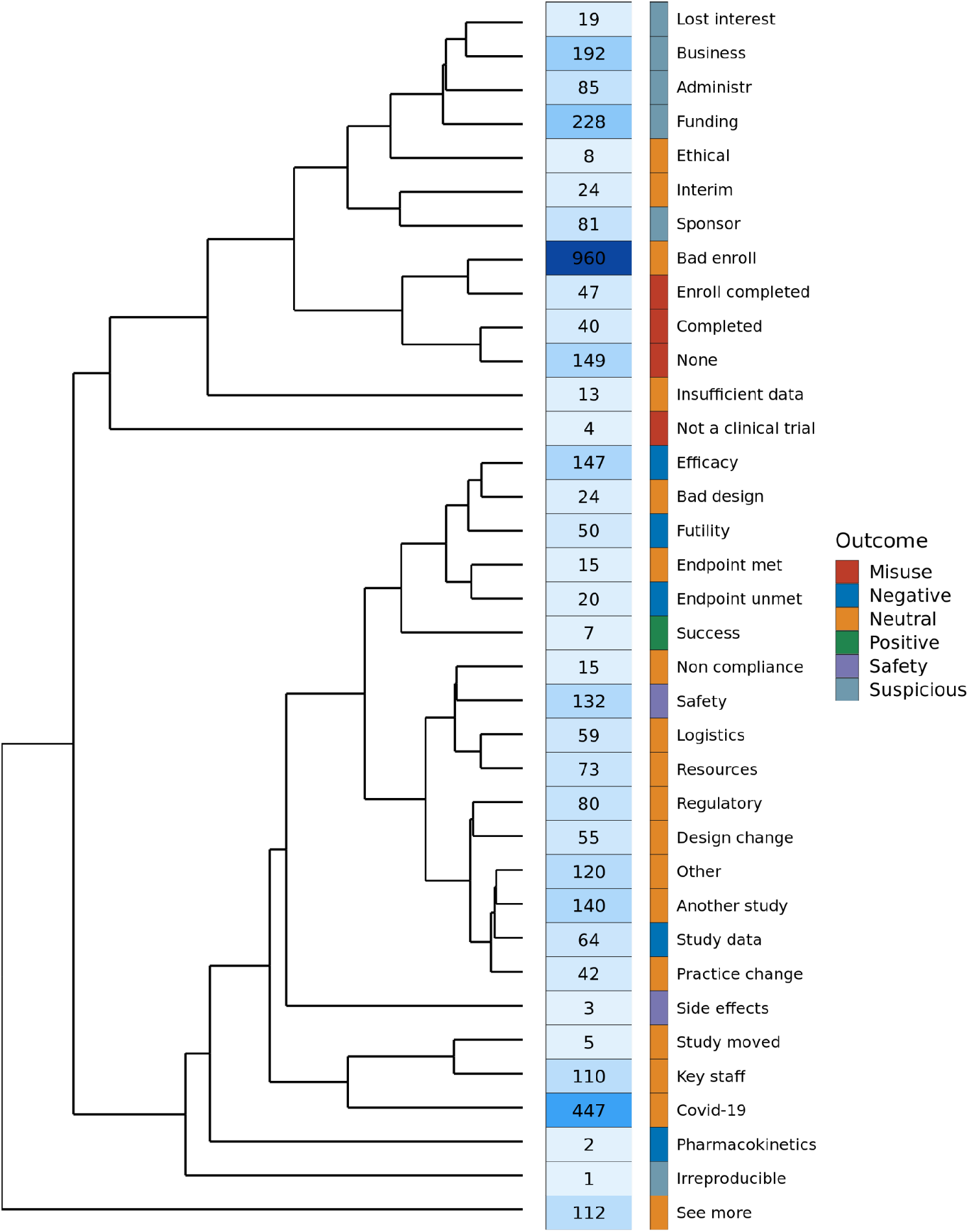
Hierarchical clustering of stop reason similarity based on curation from Pak et al. Distances were estimated as the cosine similarity of the averaged embeddings (see Supplementary Methods).

**Supplementary Figure 2.**
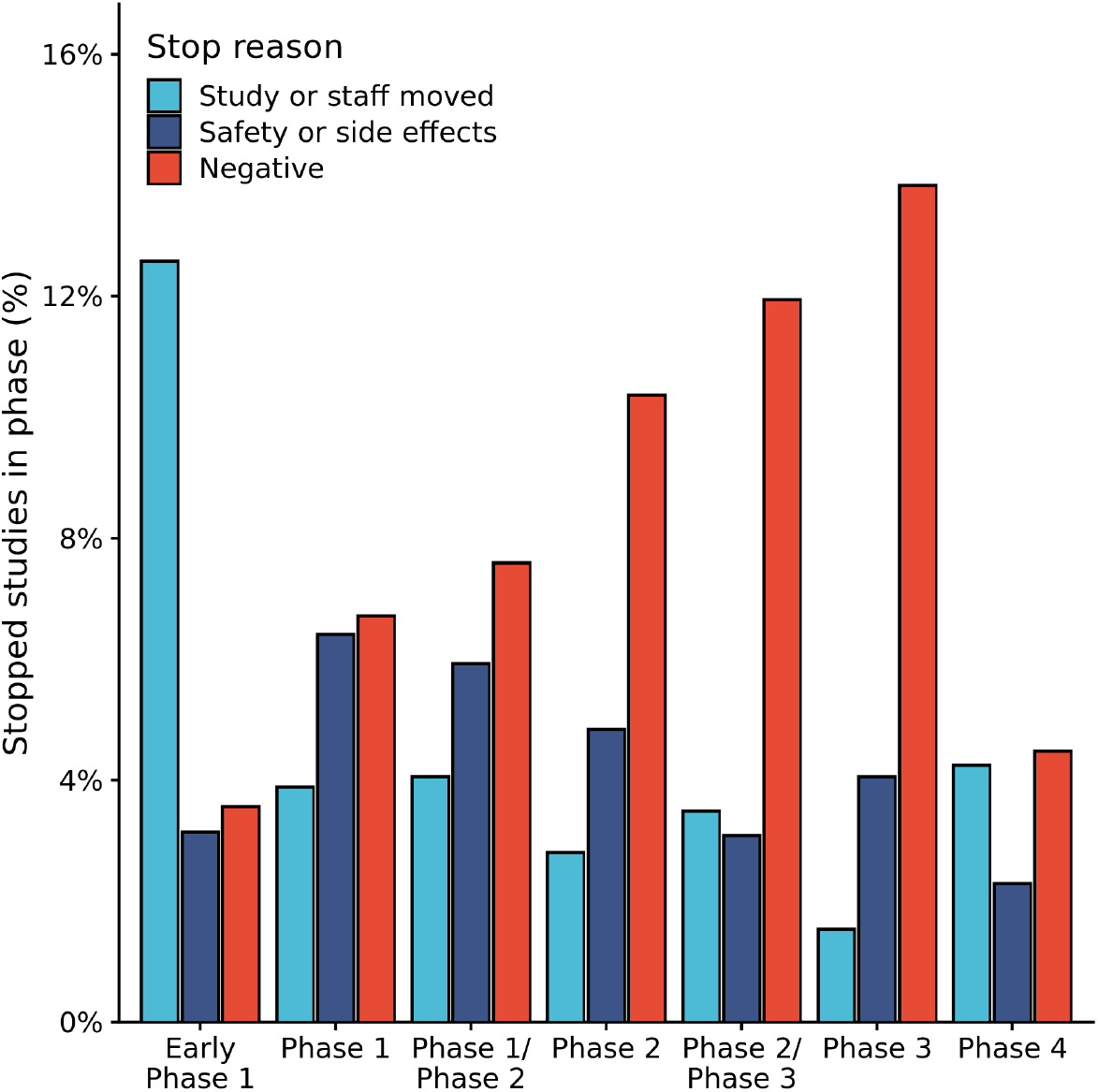
Percentage of stopped studies predicted to be due halted due to study or staff move, safety or side effects or negative reasons (e.g. efficacy) by clinical trial phase as reported by ClinicalTrials.gov.

**Supplementary Figure 3.**
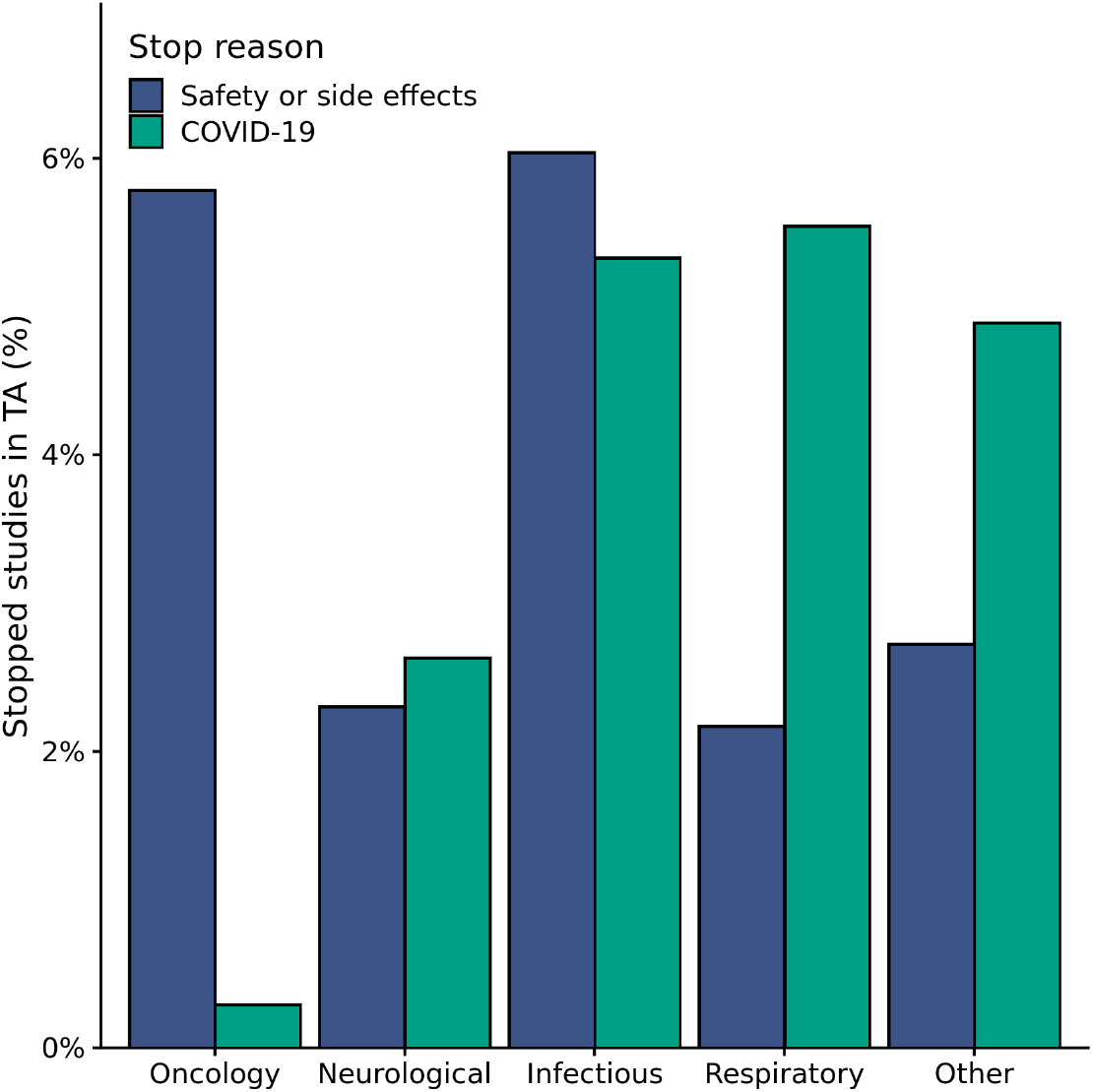
Percentage of stopped trials predicted to be halted due to Safety or side effects or the COVID-19 pandemic by predominant therapeutic area. Indications with multiple possible therapeutic areas were associated with the most severe area (e.g. Oncology).

**Supplementary Figure 4.**
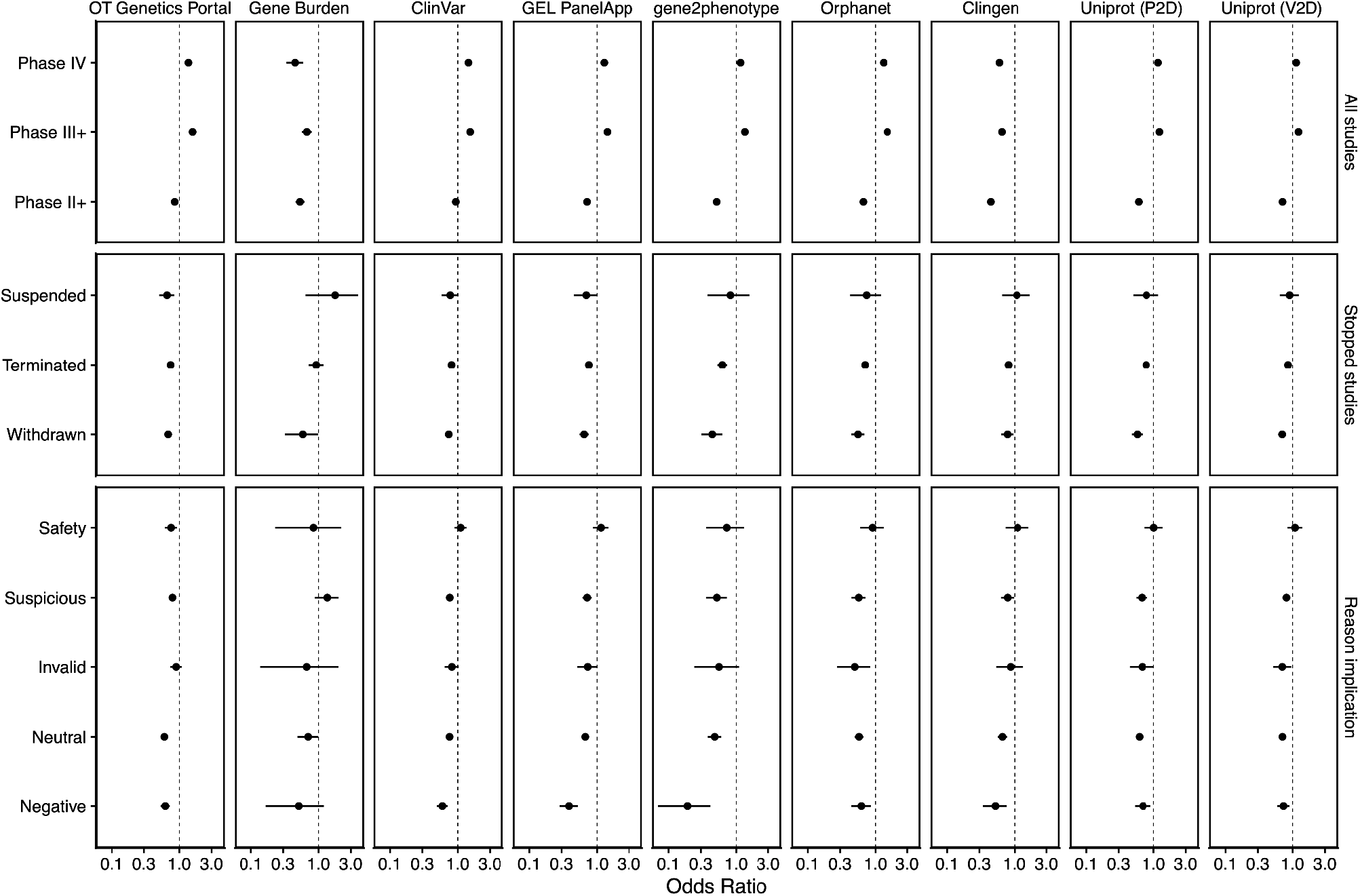
Association between the availability of genetic evidence and clinical trial outcomes by genetic data source. X-axis displays the respective odds ratio and y-axis groups the studies by phase (top row), stopped clinical trials (centre row) and stopped clinical trials split by high-level stopping reason (bottom row).

**Supplementary Figure 5.**
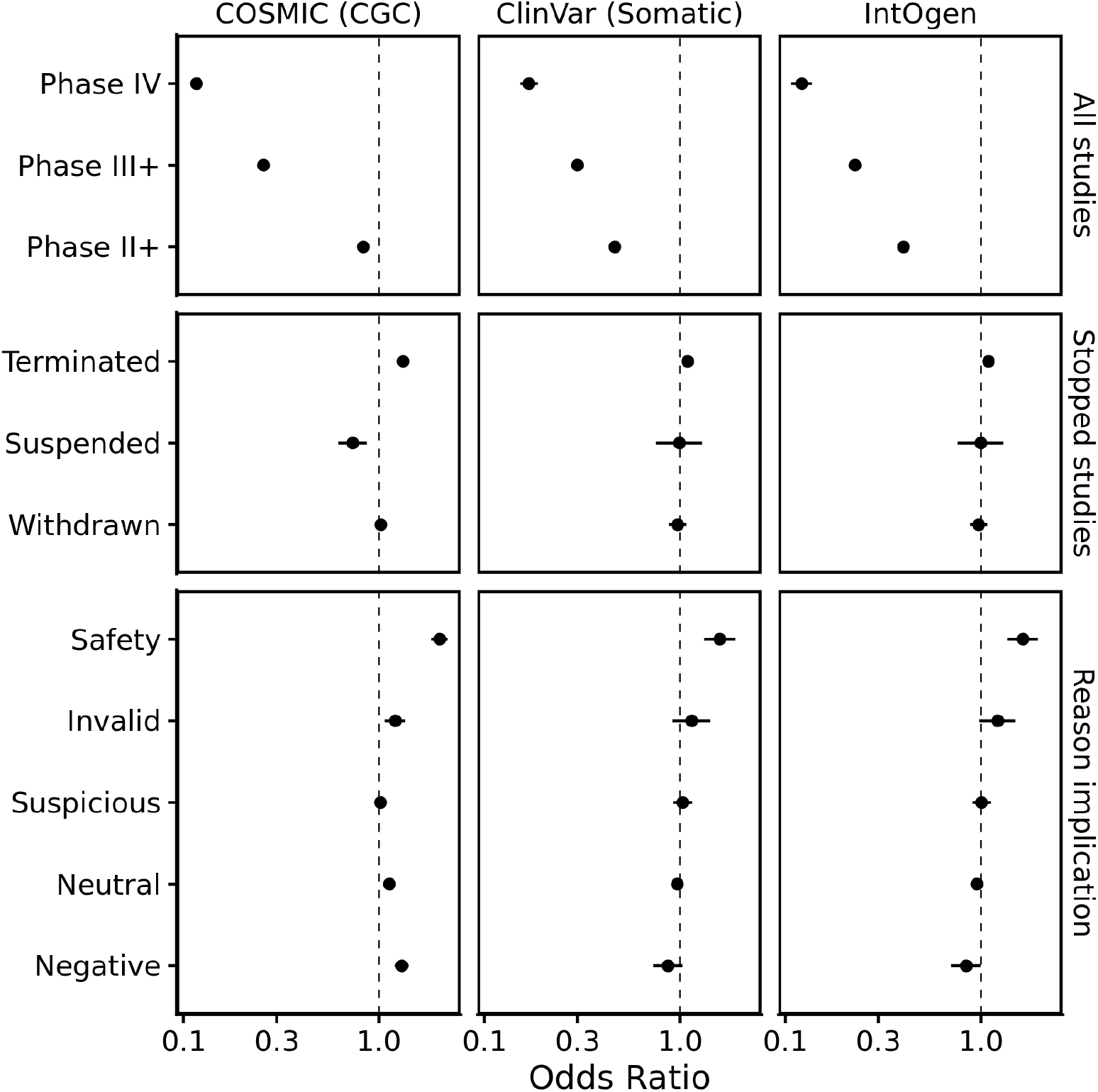
Association between the availability of genetic evidence and clinical trial outcomes by somatic data source. X-axis displays the respective odds ratio and y-axis groups the studies by phase (top row), stopped clinical trials (centre row) and stopped clinical trials split by high-level stopping reason (bottom row).

