## Supplementary Methods for "Why Clinical Trials Stop: The Role of Genetics"

Olesya Razuvayevskaya[1,2], Irene López[1,2], Ian Dunham[1,2,3] and David Ochoa[1,2]

### Redefinition of Pak *et al.* classification

In order to quantify the semantic structure of the clinical trial stop reasons, we analysed the classification of the stopped clinical trials that was developed by Pak et al. using the following strategy (Pak, Rodriguez and Roth, 2015). We trained a Long-Short Term Memory network (LSTM) to create the representations for each termination reasons and averaged the embeddings across all the examples belonging to a particular class. The class embeddings were then been used to calculate the cosine similarities among classes and visualised using an agglomerative hierarchical clustering (Supplementary Figure 1). The hierarchical representation illustrates the clusters that are semantically close to each other along with the number of examples per class and the parent category.

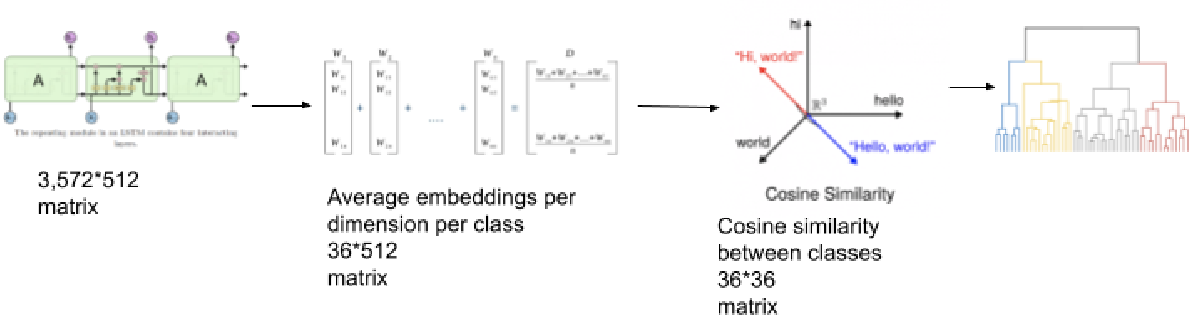


The analysis of the cosine similarities between the classes revealed that the sub-classes belonging to a certain metaclass are often clustered together as semantically similar. For example, four out of five trials belonging to the "Suspicious" sub-class were clustered together, and three out of four trials annotated as "Misuse" belong to one cluster. One of the reasons for having certain sub-classes further apart can be explained by a small number of examples, such as the "Not a clinical trial" category which only consists of four examples, or Side effects which is only represented by three examples. Another reason for why certain classes from different metaclasses are semantically similar may the multiclass nature of the dataset -- the annotators were not restricted to specifying just one sub-class for every termination reason. This resulted in having the same description corresponding to up to three different metaclasses.

Based on the analysis of the annotated stopped trials, we redefined the categories by taking into account the following factors:

- The class size (whether at least 20 instances fall into the class)
- Supervised assessment of the stop reasons to merge by clinical trial expert
- The metaclass assigned to a certain class in the original study
- The semantic similarity between the freetext descriptions

If at least three out of four criteria listed above were satisfied, we merged the subclasses into one bigger class. For example, the classes "Study Moved" and "Key Staff Left" are semantically clustered together, the categories attend to similar underlying reasons, the "Study Moved" class only consists of five examples, and both classes belong to the same parent class "Neutral".

The category called "other" consisted of very diverse reasons and often rare that can be classified into one of the given categories, with most of these reasons referring to "Invalid reason" or "No context given". We collapsed this category by removing it for the trials that had other category or categories assigned, and manually re-assigned the trials where "other" was the only class to one of the remaining categories. For example "Strategic decision unrelated to safety or efficacy" was initially classified as "Other", while it also fits "business decision", "lost interest" or "sponsor's decision" under the new "Business and administrative" category. Having this trial annotated with "Other" would be confusing for the classifier.

**List of merged categories from Pak *et al.* and resulting new category**

| **Pak *et al.* category** | **Revised category** |
|---|---|
| Lost Interest<br>Administrative reasons<br>Business decision<br>Sponsor’s decision<br>Funding | Business and administrative |
| Not a clinical trial<br>Study completed | Invalid Reason |
| Based on data in this study<br>Futility<br>Insufficient efficacy<br>Pharmacokinetics<br>Unmet endpoint | Negative |
| Safety<br>Side effects | Safety and Side Effects |
| Logistics<br>Resources | Logistics and Resources |
| Key staff left<br>Study moved | Staff or Study Moved |
| Inadequate Design<br>Change in Practice<br>Irreproducible Results<br>Non-Compliance<br>Study Design | Study Design |
| Other | - |
| Endpoint Met<br>Enrollment completed | Endpoint Met |
| None Given<br>See elsewhere | No Context |

### Classification model

We fine-tuned BERT model for the task of predicting the stop reasons on the training set of 3,571 human-annotated stopped clinical trials (Devlin *et al.*, 2018). We used a BERT uncased pre-trained model with a one-layer feed-forward classifier. The fine-tuning was performed by using the Hugging Face transformer library (Wolf *et al.*, 2019). The classifier uses 50 hidden units and the ReLu activation function.

We used the last hidden state at token "[CLS]" to retrieve a representation of the whole explanation and fed it into the classifier. We then applied "softmax" over the logits to retrieve the probabilities. The best accuracy on the validation set was achieved while training the model for 7 epochs with the batch size of 32, the learning rate of 4e-5 and Adam's optimizer with weights decay. The test set was created in a stratified manner to ensure that the relative class frequencies are taken into account in each fold of the test set.

### Human annotation study

To validate the proposed revised annotation scheme and to expand the training set, we performed a human annotation study. To perform the human annotation experiment, we extracted reasons from clinical trials deposited in ClinicalTrials.gov that were not included into the original dataset. Since the main goal of the human annotation experiment is to enrich the training data, we included as many unique trials as possible. At the same time, we also had a secondary goal of estimating the inter-annotator agreement (IAA) as an upper boundary for what the models can achieve. To combine the two goals, we randomly selected six sets of 225 unique trials with termination reasons and added 25 random trials in such a way that they overlapped between three pairs of documents. The 25 common trials were used to calculate the IAA. In addition, since the number of the annotators was odd, we generated one extra document without the overlapping trials. The annotation format consisted of descriptions and three dropdown lists next to them to select the categories from. The IAA was calculated using the Kappa statistic (K) (Cohen, 1960). If two or more annotators working independently are able to reach a substantial agreement, we can argue that the task is objective enough for the automation stage. Various schemes to assess kappa's significance have been proposed (Rietveld and van Hout, 1993). K in the range of .61 and .80 is considered substantial and K over .80 signifies an almost perfect agreement (Landis, Richard Landis and Koch, 1977).
